# Performance of ChatGPT on Chinese National Medical Licensing Examinations: A Five-Year Examination Evaluation Study for Physicians, Pharmacists and Nurses

**DOI:** 10.1101/2023.07.09.23292415

**Authors:** Hui Zong, Jiakun Li, Erman Wu, Rongrong Wu, Junyu Lu, Bairong Shen

## Abstract

**Background:** Large language models like ChatGPT have revolutionized the field of natural language processing with their capability to comprehend and generate textual content, showing great potential to play a role in medical education.

**Objective:** This study aimed to quantitatively evaluate and comprehensively analysis the performance of ChatGPT on three types of national medical examinations in China, including National Medical Licensing Examination (NMLE), National Pharmacist Licensing Examination (NPLE), and National Nurse Licensing Examination (NNLE).

**Methods:** We collected questions from Chinese NLMLE, NPLE and NNLE from year 2017 to 2021. In NMLE and NPLE, each exam consists of 4 units, while in NNLE, each exam consists of 2 units. The questions with figures, tables or chemical structure were manually identified and excluded by clinician. We applied direct instruction strategy via multiple prompts to force ChatGPT to generate the clear answer with the capability to distinguish between single-choice and multiple-choice questions.

**Results:** ChatGPT failed to pass the threshold score (0.6) in any of the three types of examinations over the five years. Specifically, in the NMLE, the highest recorded score was 0.5467, which was attained in both 2018 and 2021. In the NPLE, the highest score was 0.5599 in 2017. In the NNLE, the most impressive result was shown in 2017, with a score of 0.5897, which is also the highest score in our entire evaluation. ChatGPT’s performance showed no significant difference in different units, but significant difference in different question types. ChatGPT performed well in a range of subject areas, including clinical epidemiology, human parasitology, and dermatology, as well as in various medical topics such as molecules, health management and prevention, diagnosis and screening.

**Conclusions:** These results indicate ChatGPT failed the NMLE, NPLE and NNLE in China, spanning from year 2017 to 2021. but show great potential of large language models in medical education. In the future high-quality medical data will be required to improve the performance.

## Introduction

In the last decade, artificial intelligence (AI) technology has undergone a rapid evolution, achieving noteworthy breakthroughs in numerous fields [1, 2]. Recently, one such breakthrough that has garnered considerable attention is ChatGPT [3], an AI chatbot powered by generative pre-trained transformer (GPT) architecture, specifically GPT-3.5 with 175 billion parameters. This innovative technology is developed through human feedback reinforcement learning and trained on extensive textual data. Remarkably, ChatGPT exhibits remarkable capabilities in various tasks, including but not limited to intelligent dialogue [4], knowledge question answering[5], and text generation[6], thus showcasing unprecedented potential for further development.

In medical domain, there has been growing interest in exploration of large language models for tasks such as biomedical question answering (BioGPT [7]), and automatic dialogue generation (DialoGPT [8, 9]). Regrettably, these studies have so far demonstrated limited practical utility in clinical practice. However, ChatGPT, with its powerful language understanding and generation capabilities, showing significant potential in the fields of clinical response generation [5, 6], clinical decision support [4, 10, 11], medical education [12, 13], literature information retrieve [14], scientific writing [15-18], and beyond. Recent studies have demonstrated that ChatGPT can pass the United States Medical Licensing Exam (USMLE) [19, 20], Radiology Board-style Examination [21], UK Neurology Specialty Certificate Examination [22], and Plastic Surgery In-Service Exam [23], with results that are comparable to those of human experts. Nevertheless, Other studies have also indicated that ChatGPT failed to pass the Family Medicine Board Exam [24], and Pharmacist Qualification Examination [25]. Possible explanations for this performance difference include language and cultural differences, variations in examination content [26]. These studies highlighted the ChatGPT’s ability to comprehend the complex language used in medical contexts and its potential for use in medical education. However, current researches are limited in two aspects. Firstly, it largely focuses on the English language, and secondly, it predominantly emphasizes the physician’s examination. Additional investigation is necessary to explore the potential of ChatGPT in other non-English languages and various medical examinations, which can deliver substantial benefits for its expanded application in the medical domain.

China, with a population of over 1.4 billion, faces a significant medical burden. The provision of healthcare services involves a collaborative effort among physicians, pharmacists, and nurses who work diligently to offer the best possible care to patients. Physicians are responsible for diagnosing and treating illnesses, pharmacists ensure the appropriate medication is dispensed and administered correctly, while nurses attend to patients’ daily medical management and care service. Due to limited medical resources, medical professionals in China face immense pressure, but remain committed to providing high-quality services. The advent of ChatGPT offers a promising solution to ease this burden by delivering intelligent, efficient, and precise medical services to physicians, pharmacists, and nurses.

Medical examinations, including the Chinese National Medical Licensing Examination (NMLE), the Chinese National Pharmacist Licensing Examination (NPLE), and the Chinese National Nurse Licensing Examination (NNLE) are implemented by the government to improve professional standards, ensure medical safety and enhance healthcare services quality [27]. Through these medical examinations, the medical knowledge, clinical skills, and ethical standards mastered by medical staffs can significantly improve the quality of their services. This, in turn, can reduce the incidence of medical errors and accidents, and protect the fundamental right to health and safety of patients.

These medical licensing examinations aim to comprehensively evaluate candidate’s knowledge of medical science, clinical examination, disease diagnosis, surgical treatment, patient prognosis, policies, and regulations, among other areas. Successfully passing these examinations is a prerequisite for obtaining professional certification for physicians, pharmacists, and nurses.

In this study, we aimed to quantitatively evaluate the performance of ChatGPT on three types of national medical examinations in China, namely NMLE, NPLE and NNLE. To enhance the reliability of our findings, we meticulously collected a substantial corpus of real-world medical question-answer data from examinations conducted from the year 2017 to 2021. We also conducted a comparative analysis of the performance of different units. For cases where incorrect responses were generated, we solicited feedback from domain experts and performed thorough assessment and error analysis. Our study yields valuable insights for researchers and developers to improve large language models’ performance in the medical domain.

## Methods

### Medical examination datasets

We collected questions from Chinese NMLE, NPLE and NNLE from year 2017 to 2021. In NMLE, each exam consists of 4 units, each unit has 150 questions, for a total of 600 questions. In NPLE, each exam consists of 4 units, each unit has 120 questions, for a total of 480 questions. In NNLE, each exam consists of 2 units, each unit has 120 questions, for a total of 240 questions. Base on the requirements of the examination, a correct response rate of 60% or higher is considered to meet the passing criteria. The questions with figures, tables or chemical structure were manually identified and excluded by a clinician with five years of clinical experience.

### Model setting

We employed ChatGPT, an artificial intelligence chatbot built upon the generative pre-trained transformer technology. The official API was utilized to invoke the chatbot, with gpt-3.5-turbo as model parameter and default values for other parameters. As shown in Figure 1, the input question consisted of the background description and choices. To elicit diverse responses, we applied direct instruction strategy via prompt, such as “Please return the most correct answer”, “Only one best option can be selected”, “The correct choice is” and “This is a multiple choices question, please return the correct answer”. These prompts force the model to generate the clear answer, as well as the capability to distinguish between single-choice and multiple-choice questions.

**Figure 1.**
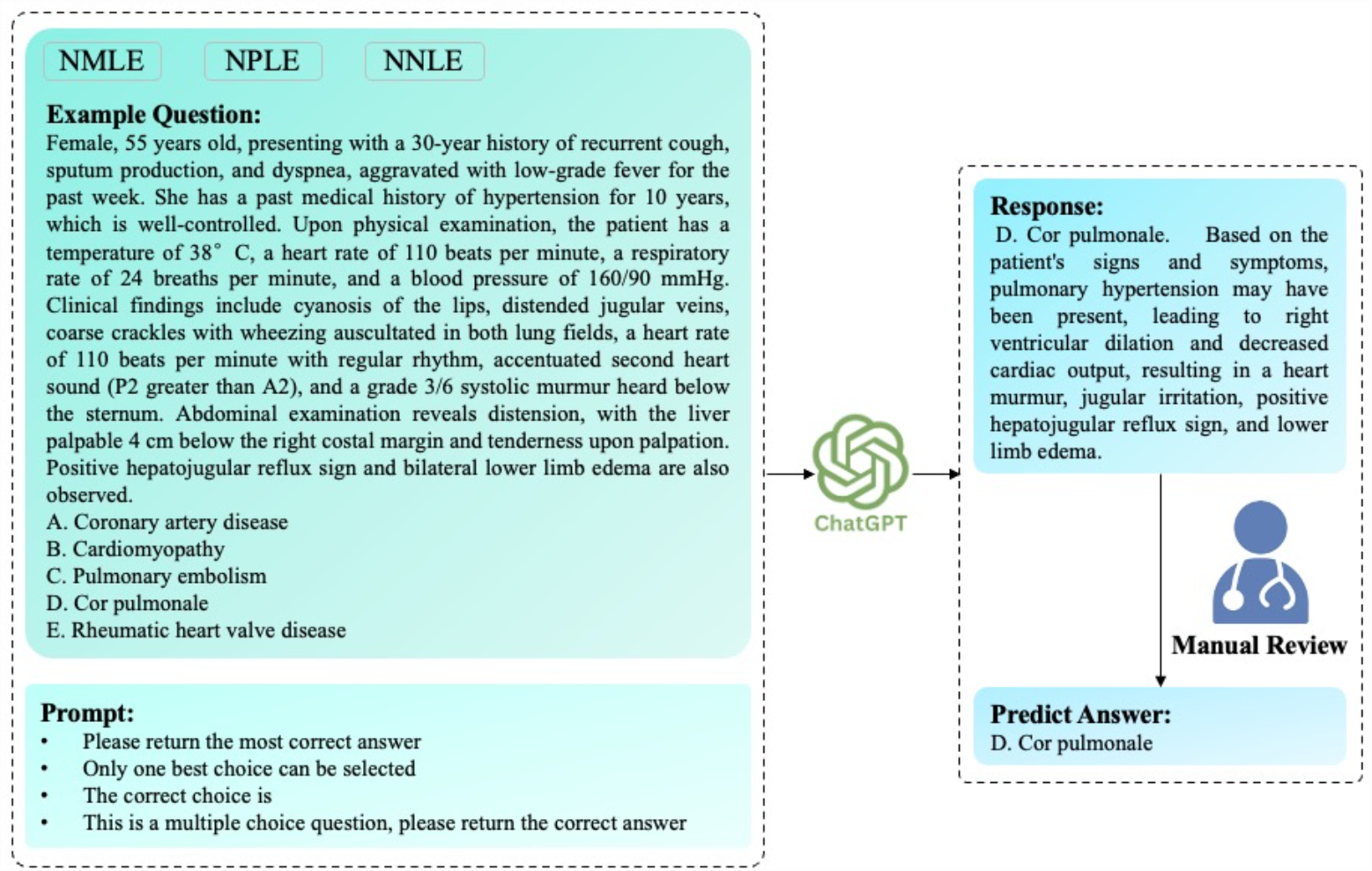
The overview of interaction with ChatGPT. The question included background description and choices from three national licensing examinations, including Chinese National Medical Licensing Examination (NMLE), National Pharmacist Licensing Examination (NPLE) and National Nurse Licensing Examination (NNLE). The prompt was designed to force a clear answer, as well as the ability to recognize single-choice or multiple-choice questions. The response of ChatGPT were manually reviewed by an experienced clinician to determine the answer. The correct answer to this question is “D. Cor pulmonale”. It should be noted that while English text was shown in the figure, the experiment itself used Chinese text as both the input and output language.

### Evaluation

For each question, the response of ChatGPT was reviewed by an experienced clinicians to determine the predict answer, which was then compared with the true answer. The score was calculated based on whether the answers match or not. A score of 1 was awarded if there is agreement between the predict answer and true answer, whereas a score of 0 was given if there is disagreement. The evaluation process has been conducted on all data sets of NMLE, NPLE and NNLE over past five years.

### Data analysis

Data process was performed in Python (version 3.9.13, Python Software Foundation) using Jupyter Notebook. Statistical analysis was performed using GraphPad Prism 9 Software. The significance of differences among groups was set at p < 0.05.

## Results

### Overall performance

As shown in Figure 2, ChatGPT failed to pass the threshold score (0.6) in any of the three types of examinations over the five years. Specifically, in the Chinese NMLE, the highest recorded score was 0.5467, which was attained in both 2018 and 2021. In the Chinese NPLE, the highest score was 0.5599 in 2017. In the Chinese NNLE, the most impressive result was shown in 2017, with a score of 0.5897, which is also the highest score in our entire evaluation. Conversely, the 2019 NPLE exam resulted in the lowest score, with a recorded value of 0.4356.

**Figure 2.**
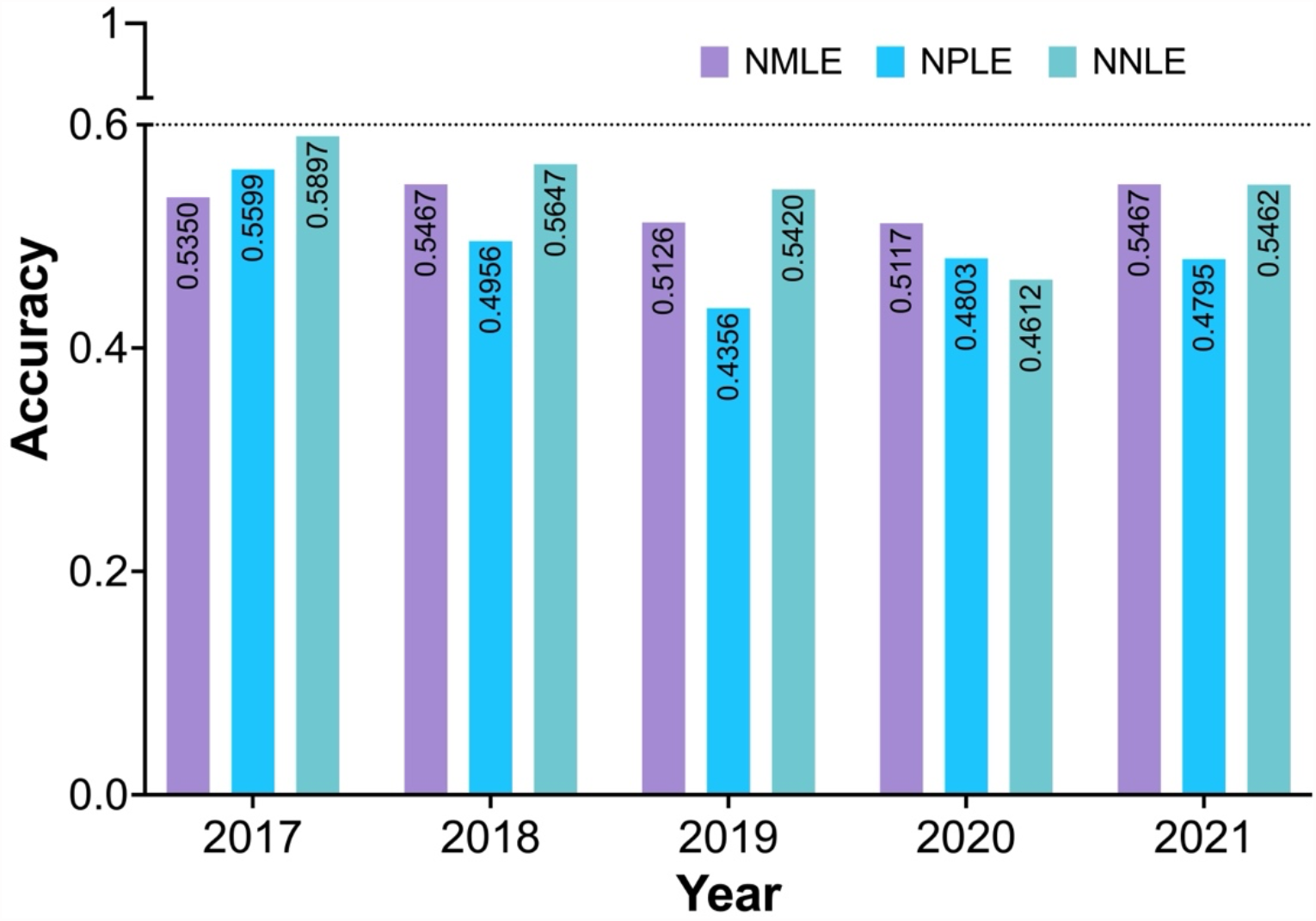
The performance of ChatGPT of three national licensing examinations over a period of five years from 2017 to 2022. The examinations included Chinese National Medical Licensing Examination (NMLE), National Pharmacist Licensing Examination (NPLE) and National Nurse Licensing Examination (NNLE).

### Detailed performance

The score of each unit in the Chinese NMLE is shown in Table 1. The performance of ChatGPT has varied across different units and years. In 2017 and 2020, ChatGPT performed best in Unit 2. In 2018 and 2019, ChatGPT performed best in Unit 1. In the 2021, ChatGPT performed best in both Unit 2 and Unit 3. In 2018 and 2021, ChatGPT correctly answered 328 out of 600 questions. This is because the complexity and difficulty of question in each unit were vary from year to year. On average, ChatGPT achieved the highest score in Unit 2 (84.6), followed by Unit 1 (79.8), Unit 3 (78.2), Unit 4 (75.4).

**Table 1.** The score of each unit in Chinese National Medical Licensing Examination.

| Year | 2017 | 2018 | 2019 | 2020 | 2021 | Average |
| --- | --- | --- | --- | --- | --- | --- |
| Unit1 score | 81 | 87 | 84 | 71 | 76 | 79.8 |
| Unit2 score | 96 | 83 | 75 | 83 | 86 | 84.6 |
| Unit3 score | 79 | 79 | 72 | 75 | 86 | 78.2 |
| Unit4 score | 65 | 79 | 75 | 78 | 80 | 75.4 |
| Total score | 321 | 328 | 306 | 307 | 328 | 318 |
| Questions | 600 | 600 | 597 | 600 | 600 | - |
| Accuracy | 53.50% | 54.67% | 51.26% | 51.17% | 54.67% | 53.05% |

The score of each unit in the Chinese NPLE is shown in Table 2. In NPLE, each unit has 120 questions, and each exam has 480 questions. We identified and removed the questions included figures, tables or chemical structure. Such questions appeared the most in 2018 (30), followed by 2020 (22), 2017 (21), 2021 (17) and 2019 (14). On average, the ChatGPT performed best in Unit 4 (62), followed by Unit 2 (58), Unit 3 (53) and Unit 1(52). In the year 2017, ChatGPT achieved highest score, and correctly answered 257 out of 459 questions.

**Table 2.** The score of each unit in Chinese National Pharmacist Licensing Examination.

| Year | 2017 | 2018 | 2019 | 2020 | 2021 | Average |
| --- | --- | --- | --- | --- | --- | --- |
| Unit1 score | 57 | 55 | 51 | 48 | 49 | 52 |
| Unit2 score | 63 | 54 | 52 | 60 | 61 | 58 |
| Unit3 score | 60 | 56 | 49 | 53 | 47 | 53 |
| Unit4 score | 77 | 58 | 51 | 59 | 65 | 62 |
| Total score | 257 | 223 | 203 | 220 | 222 | 225 |
| Questions | 459 | 450 | 466 | 458 | 463 | - |
| Accuracy | 55.99% | 49.56% | 43.56% | 48.03% | 47.95% | 49.02% |

The Table 3 shown the detailed score of each unit of Chinese NNLE. There are totally 26 questions included figures, tables or chemical structure were removed. In 2017 and 2018, ChatGPT performed better in Unit2 than Unit1. Conversely, in 2019, 2020 and 2021, ChatGPT performed better in Unit1 than Unit2. On average, ChatGPT’s performance of the two units had no noticeable difference.

**Table 3.** The score of each unit in Chinese National Nurse Licensing Examination.

| Year | 2017 | 2018 | 2019 | 2020 | 2021 | Average |
| --- | --- | --- | --- | --- | --- | --- |
| Unit1 score | 65 | 57 | 72 | 54 | 67 | 63 |
| Unit2 score | 73 | 74 | 57 | 53 | 63 | 64 |
| Total score | 138 | 131 | 129 | 107 | 130 | 127 |
| Questions | 234 | 232 | 238 | 232 | 238 | - |
| Accuracy | 58.97% | 56.47% | 54.2% | 46.12% | 54.62% | 54.08% |

In comparison, ChatGPT exhibited better proficiency in NNLE (54.08%), with NMLE (53.05%) and NPLE (49.02%) following behind. The result corresponds to the complexity and difficulty of the exam questions.

### Performance on different units and question types

Figure 3 demonstrated the comparative analysis of ChatGPT’s performance differences across units and question types. The results shown there was no significant difference in across different units in NMLE (Figure 3A), NPLE (Figure 3B), and NNLE (Figure 3C). However, in the case of NPLE (Figure 3D), ChatGPT demonstrated higher performance in single-choice questions compared to multiple-choice questions, with a highly statistical difference (p < 0.0001).

**Figure 3.**
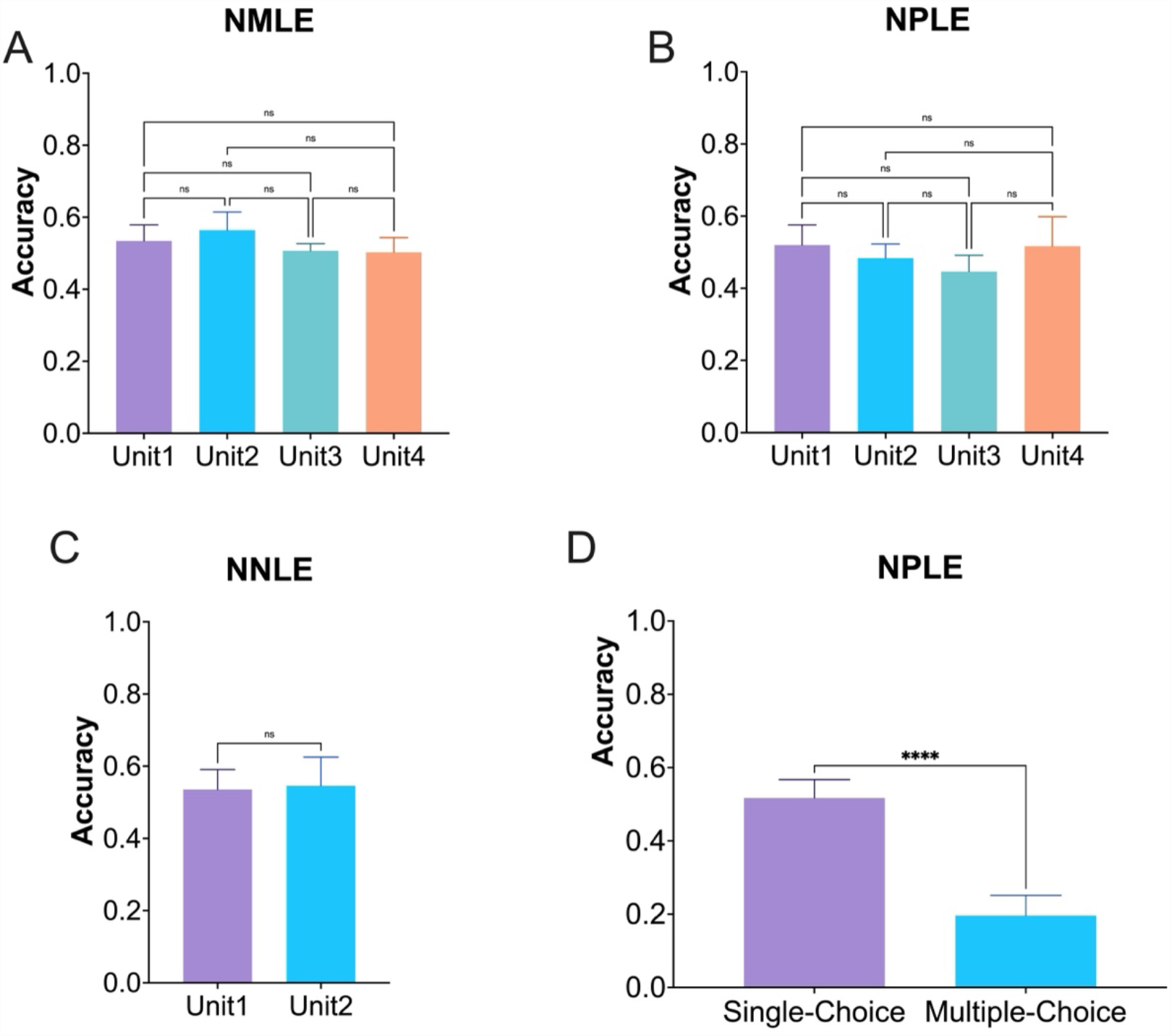
The performance of ChatGPT on different units and question types. For different units, there were no significant difference among (A) Chinese National Medical Licensing Examination (NMLE), (B) National Pharmacist Licensing Examination (NPLE), and (C) National Nurse Licensing Examination (NPLE). (D) However, ChatGPT demonstrated higher performance in single-choice questions than multiple-choice questions with a highly significant difference (ns, no significant difference, ****p < 0.0001).

### Performance on different subjects and topics

To better understand why ChatGPT failed in the Chinese medical examination, we took the 2021 NMLE exam as an example, and labeled the medical subjects and topics for each question (Figure 4). The result revealed that ChatGPT excelled in clinical epidemiology, human parasitology, and dermatology, with all questions answered correctly. However, the model faltered in subjects such as pathology, pathophysiology, public health regulations, physiology, and anatomy, with a correct rate of less than 50%. Additionally, we observed that ChatGPT performed admirably in topics related to molecule, health management and prevention, diagnosis and screening, but its performance was lackluster in topics such as clinical manifestations, indicator values, structural location, cell, and tissue. Interestingly, we found no significant difference in performance on case-based questions and non-case-based questions. The result provides deep insight into the strengths and weaknesses of ChatGPT in medical examinations, and pave the way for future research to improve the model’s capabilities in this domain.

**Figure 4.**
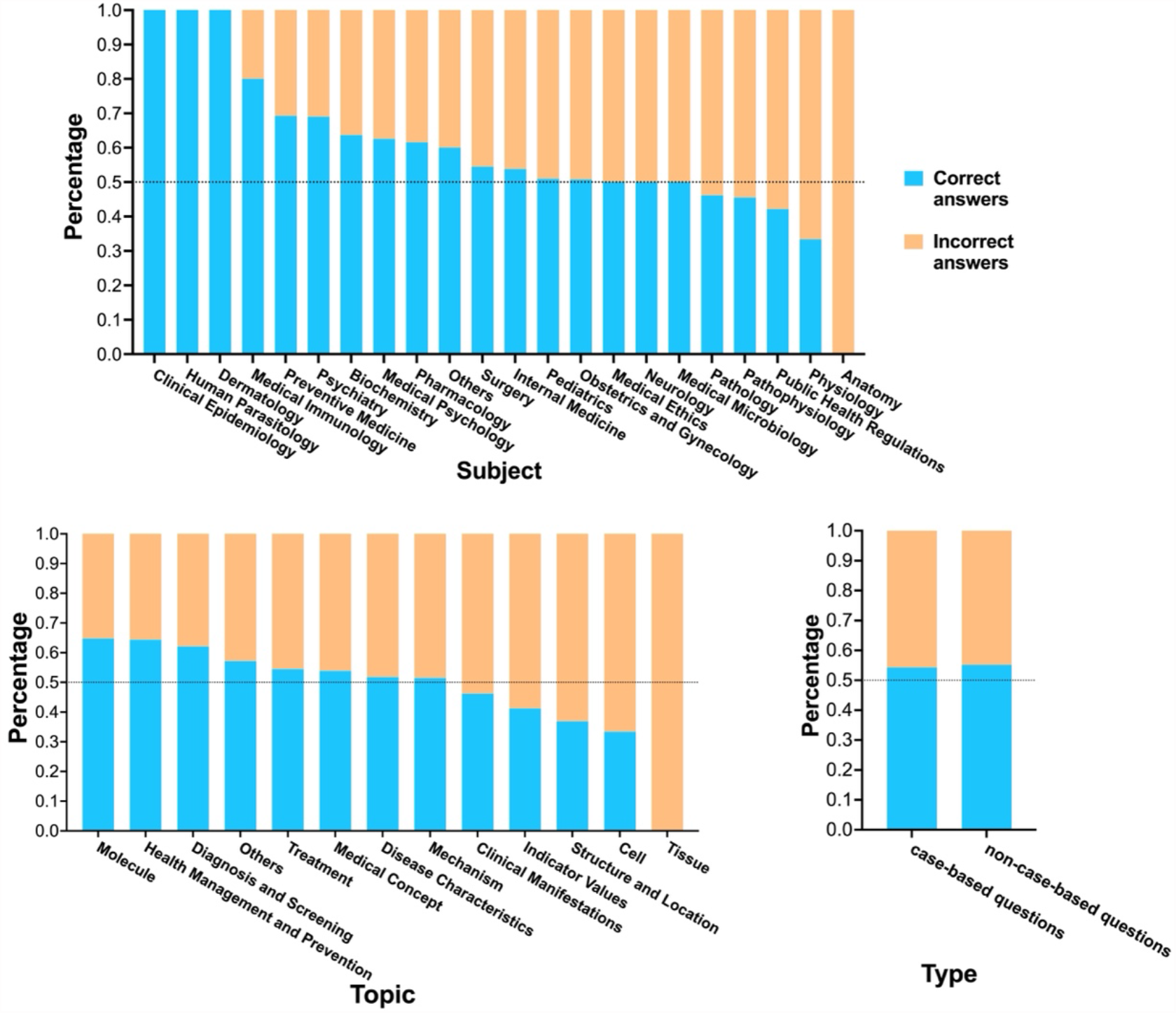
The performance of ChatGPT on different subjects, topics and types of questions in the 2021 NMLE exam.

## Discussion

In this study, we evaluated the performance of ChatGPT, an artificial intelligence chatbot, in answering medical exam questions from Chinese NMLE, NPLE, and NNLE from year 2017 to 2021.

### ChatGPT failed NNLE, NMLE and NPLE in China

The results of our study revealed that ChatGPT was unsuccessful in meeting the requirements of the three primary medical licensure assessments namely, NMLE, NPLE and NNLE in China, spanning from 2017 to 2021. There are several possible reasons for this.

Firstly, According to OpenAI, ChatGPT has been trained with the vast majority of the data is in English, with only a small amount of data in other languages, like Chinese. More richer training dataset allows the model to learn more knowledge. Recent studies shown that ChatGPT passed United States Medical Licensing Examination [19, 20]. However, it failed to pass the Taiwanese Pharmacist Licensing Examination [25], and performed worse than medical students on Korean-based parasitology examination [28]. These findings suggest that ChatGPT may require additional training data in non-English languages to enhance its performance in non-English medical exams.

Secondly, there are differences in medical policies, regulations, and management agencies across countries with different languages or cultures. In the Chinese NMLE, some questions relate to healthcare policies, while in the Chinese NPLE, the entire unit 4 is officially designated as pharmaceutical management and regulations. These questions cover topics such as drug production, market circulation, pharmaceutical management, and legal regulations. The aim is to assess awareness and compliance ability in clinical practice. Generally, these questions are relatively short in length and clear in meaning. While ChatGPT has acquired a wealth of knowledge on healthcare policies from English-speaking countries due to its extensive English dataset, it may encounter difficulties in correctly understanding the healthcare policies of non-English-speaking countries, leading to erroneous responses to related questions. Additionally, healthcare policies undergo regular updates over time, making such questions more challenging.

Thirdly, in some questions, the task requires reading the question and selecting the most suitable answer from five given choices. However, there may be more than one choice that can answer the question, and in such cases, ChatGPT may generate multiple answers, including the correct answer. As this is a single-choice question, ChatGPT is forced to select a single choice as answer, which can limit its content comprehension ability and lead to incorrect answer.

### The potential of large language model in medical education

As a significant milestone in the development of artificial intelligence, ChatGPT driven by large language model has powerful capability in language understanding and content generation. With its remarkable potential, ChatGPT could be a valuable resource in acquiring medical knowledge and learning clinical skills for students, and serve as an informative assistant in preparing teaching materials and evaluating course projects for teachers.

In our study, ChatGPT has achieved an accuracy rate of over 50% in most of the exams, indicating a significant potential for ChatGPT in medical education. Previous study shown in the Chinese Rural General Medical Licensing Examination, only 55% of students were able to pass the written examination [29]. In China, the significant healthcare burden necessitates a vast number of licensed clinical staff and healthcare providers. However, the rigorous examinations lead to low pass rates, exacerbating the shortage of licensed practitioners, especially in rural areas. The large language model presents a promising avenue for enhancing medical education and advancing healthcare reform, with the potential to reduce medical burden.

Finally, the advancement of artificial intelligence (AI), specifically large language models, in medical education needs public benchmarking datasets and fair evaluation metrics for performance assessment. There is also a need to interact with human experts from multiple dimensions and obtain continuous feedback. In addition, the use of such model must also consider data privacy, cognitive bias, and comply with regulations.

### Limitations

Our study has some limitations. First, the questions of China NMLE, NPLE, and NNLE are all multiple-choice format. While this format meets our study purposes, it did not fully showcase the content generation capabilities of ChatGPT. In the future, it would be beneficial to include more open-ended questions. Second, we evaluated the performance of ChatGPT in medical examinations with zero-shot learning. However, better performance may be achieved by incorporating knowledge-enhanced training methods. Third, the different variations of prompt may impact ChatGPT’s response, leading to diverse answers. Therefore, it is imperative to develop innovative techniques that can generate more consistent and trustworthy responses in the future. Finally, further investigation is needed to determine the underlying factors contributing to this substandard performance, and to explore broader application of ChatGPT in medical education and clinical decision-making support.

## Conclusion

In conclusion, we evaluated the performance of ChatGPT on three types of national medical examinations in China, including NMLE, NPLE, and NNLE from year 2017 to 2021. The results indicated ChatGPT failed to meet the official pass criteria of 60% correct response rate in any of the three types of examinations over the five years. The performance of ChatGPT varied across different units and years, with the highest score achieved in NNLE of year 2017. ChatGPT exhibited relatively better proficiency in NNLE, with NMLE and NPLE following closely behind. ChatGPT performed well in a range of subject areas, including clinical epidemiology, human parasitology, and dermatology, as well as in various medical topics such as molecules, health management and prevention, diagnosis and screening.

## Data Availability

The data analyzed and reported in this study are available from the authors upon reasonable request.

## Authors’ Contributions

HZ, JL, EW, RW, JL and BS involved in the study conceptualization. JL collected and preprocessed the data. HZ conducted data analysis, results interpretation and manuscript preparation. HZ, JL EW, RW, JL and BS contributed to the review and editing of the manuscript. BS supervised the study. All authors read and approved the final manuscript.

## Acknowledgments

We thank OpenAI, which allows free access to ChatGPT.

